# Detection of Type 2 Diabetes from 20-second Speech Recordings: A Large-Scale Validation Study

**DOI:** 10.64898/2026.03.16.26348468

**Authors:** Elisa Brann, Roseline Polle, Giedrė Čepukaitytė, Alexandra L. Georgescu, Owen Parsons, Barbora de Courten, Emilia Molimpakis, Stefano Goria

## Abstract

Timely and accessible screening for type 2 diabetes (T2D) remains a global priority, given the large number of cases that go undetected. Here, we present the largest known real-world validation study for a speech-based T2D prediction model, trained on speech data from over 21,000 individuals, that works on features extracted from 20-second speech recordings. The model was evaluated in two stages: 1) Against self-reported diagnoses in 7,319 English-speaking participants using AUC, and 2) Against HbA1c blood tests in a subset of 801 participants drawn from the latter cohort. Performance was also compared against QDiabetes and in the presence of key confounding variables. The model demonstrated clinically useful predictive capacity on self-reported data (AUC = 0.80 ± 0.03), approaching QDiabetes (AUC = 0.86 ± 0.03). It was robust to most demographic confounds (e.g., age and sex) and medication use, with reduced performance in the presence of comorbidities (e.g., cardiovascular disease and hypertension). At diabetes threshold of HbA1c ***≥***48 mmol/mol, the model achieved an AUC of 0.75 (±0.07). This biomarker-validated speech-based tool demonstrates potential to complement existing methods through accessible, scalable screening requiring only a 20-second speech sample.

## 1 Introduction

Type 2 diabetes (T2D) affects an estimated 7% of UK adults, with approximately 30%, or around one million cases, remaining undiagnosed[1]. While early detection is critical for preventing complications and reducing healthcare costs, current screening approaches are largely opportunistic or rely on time-intensive appointments that face significant barriers. For example, the NHS Health Check offers UK adults aged 40-74 a 20-30-minute in-person assessment of lifestyle, family history, and clinical measurements, including blood pressure, cholesterol, and body mass index (BMI), to calculate their risk of developing T2D and other chronic conditions. Although the programme has improved disease detection[2], uptake remains limited, with only 40.4% of eligible adults attending as of 2025[3]. Some of the reasons behind low uptake include aversion to preventive care, competing priorities and difficulty accessing GP services[2]. Both screening access and T2D prevalence also vary substantially across demographic groups[1]. Remote, non-invasive screening tools that require minimal time commitment from both patients and GPs may improve detection rates and extend the reach to populations currently underserved by traditional healthcare pathways.

Speech-based tools offer particular promise as an accessible, scalable and low-burden approach to early-stage screening. Speech contains rich linguistic and paralinguistic signals reflecting both physical and mental health[4]. Availability of everyday digital devices, including smartphones, facilitates large-scale collection of audio recordings that capture this information[5–7]. Patterns in speech signals, including subtle characteristics that may be imperceptible to conventional assessment, can in turn be extracted using machine learning approaches, enabling automated screening from speech recordings.

However, large-scale studies leveraging speech and voice data for disease prediction remain scarce, particularly those capturing the demographic diversity and medical complexity of real-world populations. One notable example, focused on depression and anxiety, gathered data from over 30,000 participants[7]. Models evaluated on a separate dataset from 2,431 participants predicted symptoms with an area under the curve (AUC) of 0.84[7]. Given the accessibility of speech-based assessments, a model of equivalent accuracy deployed at population scale could transform existing diabetes screening, reaching individuals who would otherwise miss out on conventional services and increasing uptake of the more time- and resource-intensive tests by those who actually need it.

T2D affects voice through multiple physiological pathways, with the extent of vocal changes generally corresponding to disease progression. Individuals with diabetes show a higher incidence of noticeable voice problems such as vocal straining, hoarseness, and significantly shorter maximum phonation time compared to healthy controls[8–10]. These more pronounced, perceptible symptoms are associated with markers of advanced disease, specifically, poor glycemic control and the presence of neuropathy. However, to date, research has largely focused on long-term cumulative damage rather than tracking voice changes across disease stages or investigating the subtler vocal alterations that may occur earlier in disease development[11].

Initial studies demonstrate the potential of speech-based approaches for T2D detection[12–17]. For example, in a large study of 3,129 participants, Guo and colleagues[12] found that models trained on paralinguistic features outperformed those that relied on clinical variables, achieving AUCs of *>*80%, with jitter and shimmer as important paralinguistic predictors. However, given the tightly controlled conditions under which data for this study were collected, including the quiet setting and professional recording equipment[12], these findings may not generalise to real-world environments. Studies by Kaufman and colleagues[13] and Elbéji and colleagues[14] achieved T2D detection accuracies of 70-75% using smartphone recordings. They identified significant vocal changes in T2D patients related to pitch, intensity, and vocal perturbations[13, 14]. However, these studies were constrained by modest sample sizes (n=267 and 607, respectively) and while both attempted to incorporate diversity into recruitment strategies[13, 14], replication in larger, population-representative cohorts is required before clinical translation to avoid amplifying preexisting disparities in early screening programmes[1, 18, 19].

The present study addresses this critical gap by validating a speech-based T2D detection model trained on more than 21,000 participants—the largest known training dataset for this task—against the largest and most comprehensive real-world validation dataset to date, comprising over 7,000 unique users, more than 12 times larger than previous real-world studies. Together with extensive personal and health information collected as part of the study, this scale enables robust assessment of performance across diverse demographic and clinical subgroups, in the presence of key confounding factors and against established screening methods. Extending previous studies in scale[12–15], we also incorporated clinical validation through haemoglobin A1c (HbA1c) testing. This allowed us to quantify changes in predictive performance from mild (prediabetes) to severe (diabetes) disease stages—a step not undertaken in previous voice-based T2D studies.

Our investigation pursued three primary objectives. First, we aimed to evaluate the speech-based model’s ability to predict self-reported diabetes status. The rich demographic and health data collected enabled stratified analysis by typical T2D risk factors, such as age, sex, ethnicity, diabetes medication use, namely, no medication or any medication, and comorbid conditions frequently associated with T2D, including cardiovascular disease (CVD), hypertension, obesity (BMI *≥* 30) and chronic kidney disease (CKD). This comprehensive approach allowed us to identify where voice-based screening performs optimally and where additional refinement may be needed.

Second, to validate model predictions beyond self-reported diagnoses, we distributed HbA1c blood test kits to over 800 participants, making this the only known large-scale, fully remote speech-based diabetes study with concurrent blood sampling, with prior studies utilising health records or self-report to obtain HbA1c levels[12–15]. Participants in this sample were drawn from the initial cohort stratified by predicted diabetes risk. By linking speech-based risk predictions to objective biomarker measurements obtained within three months of the speech samples, we evaluated the model’s ability to detect T2D across the disease continuum, in prediabetes and diabetes stages, providing validation against a clinical gold standard independent of self-report.

Third, we compared our speech model against QDiabetes, the National Institute for Health and Care Excellence (NICE)-recommended tool that uses demographic and clinical variables to estimate 10-year T2D risk[20, 21]. In practice, no systematic non-invasive screening pathway exists for T2D in the UK: current detection is largely opportunistic, relying on blood tests taken incidentally during GP appointments. While QDiabetes is recommended for risk identification[21], it is underutilised[2, 3], and, importantly, predicts future risk rather than current disease status[20]. In the absence of a true non-invasive screening comparator, QDiabetes represents the most relevant available benchmark. By directly comparing performance, we assessed whether analysis of 20-second speech segments offers comparable detection capabilities to existing but under-exploited screening tools based on risk scores computed from demographic and clinical variables and requiring greater time commitment.

## 2 Methods

### 2.1 Study Design

This study employed a two-stage approach to validate a voice-based T2D screening tool. Stage 1 evaluated the speech-model performance on a prospectively collected cohort of 7,319 UK adults who completed voice recordings and health questionnaires, with predictions compared against self-reported T2D diagnoses. This cohort was distinct from the model’s training dataset (n = 21,129 participants; see Section 2.4). Stage 2 validated model predictions against HbA1c blood biomarkers in a stratified subset of 801 UK adults, a subset of the Stage 1 participant cohort, selected to ensure representation across model-predicted levels of risk of T2D. This design, represented in Figure 1, enabled evaluation of model discrimination against both patient-reported diagnoses and objective clinical biomarkers. This study was retrospectively registered on ClinicalTrials.gov (NCT07421921) on 11/27/2025.

**Fig. 1.**
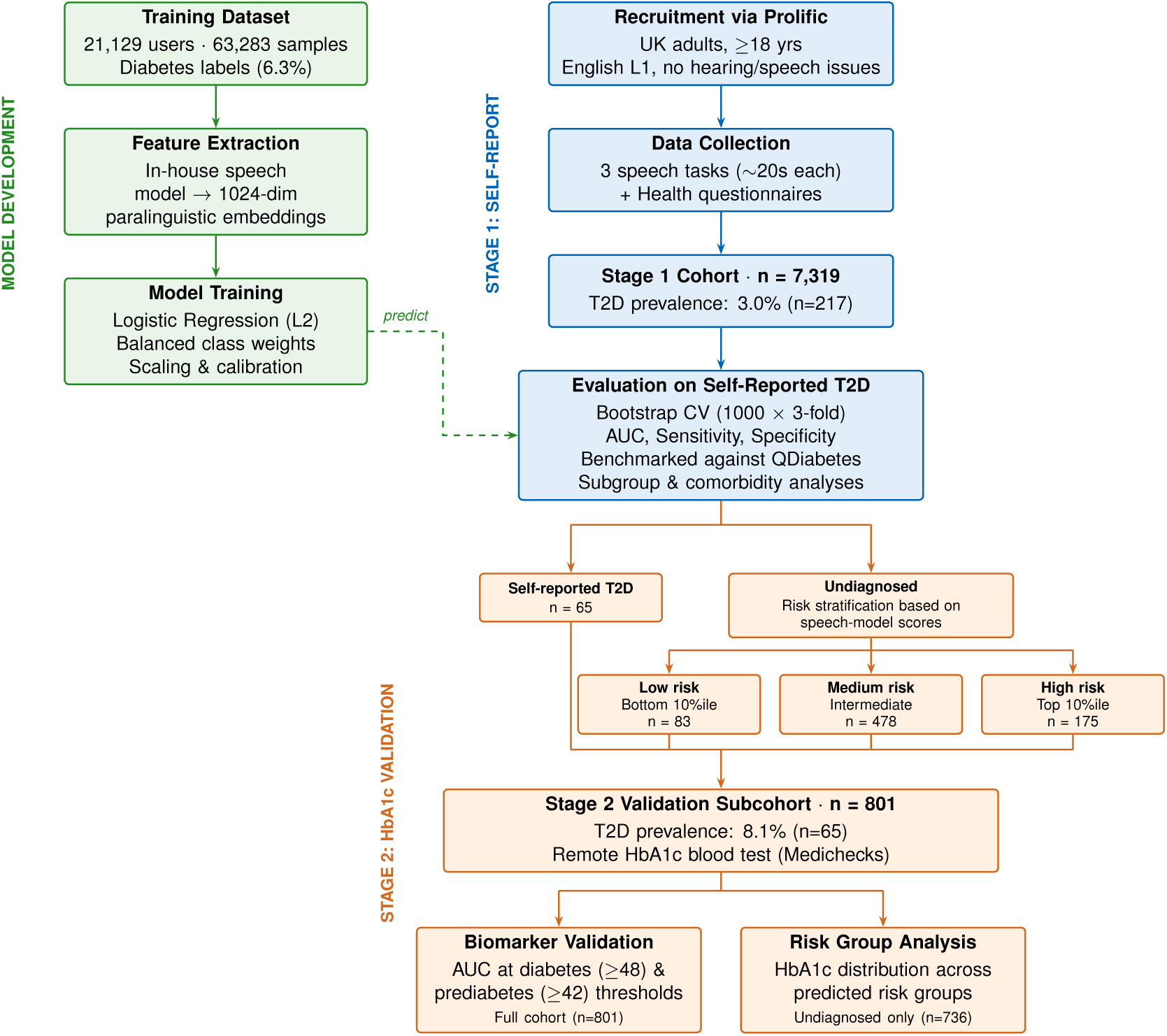
Study design. The speech model was developed using a proprietary dataset of 63,283 voice samples from 21,129 unique English-speaking participants residing in the UK or US. The model was evaluated in two stages: Stage 1 included a separate cohort of 7,319 English-speaking participants from the UK who provided self-report data; Stage 2 involved a sub-cohort of Stage 1 who completed home HbA1c tests for validation against gold-standard. This subset of participants included both individuals with self-reported type 2 diabetes (T2D) and without self-reported diabetes diagnosis (Undiagnosed). Undiagnosed participants were selected for Stage 2 based on model-predicted diabetes risk.

### 2.2 Participants

Participants were recruited using an online research participation platform (Prolific) and were required to be 18 years or older, speak English as a first language, have no language difficulties, have normal or corrected to normal eye-sight, no hearing impairments, and be a current resident in the UK.

### 2.3 Ethical Considerations

The ethical review process for this validation study was led by the University of Portsmouth Research Ethics Committee (25/ETHICS/011), with a favourable opinion granted on August 19, 2025. An amendment to add secondary data analysis for model training was approved on March 20, 2026. All participants gave written informed consent and were compensated for their time via Prolific. Individuals who completed the HbA1c test were also provided with access to a written personalised doctor’s report detailing the results of their test and follow-on care guidance. All study procedures were performed in accordance with the Declaration of Helsinki.

### 2.4 Model Development and Training

#### Training data

The training data processing and model development pipeline is illustrated in Figure 1 (top left). The model was developed using a proprietary dataset of 63,283 voice samples from 21,129 unique English-speaking participants whose country of residence was either the United Kingdom or the United States, collected via the thymia research platform[22]. Recordings comprised a reading-aloud task and two free-speech tasks. Alongside speech data, participants completed a general health questionnaire that asked whether they had been diagnosed with diabetes of *any* type. As the original data collection was not designed around Type 2 diabetes specifically, the questionnaire did not distinguish between diabetes types; we refer to this as the *noisy* or *mixed-type* diabetes label, with an estimated prevalence of 6.3% (1,318 participants). Of these, an estimated 80-90% are type 2, consistent with UK and US population estimates[23, 24]. The model was subsequently evaluated in two stages using *clean* Type 2 diabetes labels: first against self-reported T2D status from a targeted health questionnaire (Stage 1; Section 2.5.1), and then against HbA1c-derived labels from remote blood testing (Stage 2; Section 2.5.2).

#### Feature extraction

Voice recordings were preprocessed through an in-house pipeline involving resampling to 16kHz and trimming silences to produce segments of 10-20s. Paralinguistic 1024-dimensional features were extracted from trimmed audio segments using an in-house speech model based on TRILLsson5[25]. To balance data quality with real-world generalisability, minimal pre-processing was applied. Samples were excluded only if insufficient speech was present to reach the 10-second threshold or recording quality was inadequate for automated transcription. Here we use an automated-transcription tool (Deepgram base model) only as a quality control tool, as well as to trim silences. These criteria excluded approximately 5% of collected samples. We additionally excluded samples missing questionnaire data required for QDiabetes comparison (see Section 2.6.2) calculation (e.g., weight), corresponding to 7% of collected samples. Due to overlap between these exclusion criteria, 10% of samples were dropped in total. All reported numbers and results use this filtered dataset.

#### Prediction model

A logistic regression classifier with L2 regularisation (*C* = 0.001) was trained on the 1024-dimensional embeddings using the lbfgs solver and balanced class weights to account for the low prevalence of diabetes in the training population. A standard scaler was applied to normalise the input features. The model was calibrated using Platt scaling (sigmoid method) via 3-fold cross-validation on the training set, so that among individuals assigned a predicted probability of X%, approximately X% are expected to have the condition.

### 2.5 Data Collection

#### 2.5.1 Stage 1: Voice and Self Reported Health Information

Individuals completed the same speech activities as described in Section 2.4. The reading out loud task involved a standard text commonly used as a speech elicitation task due to its phonetic range (the Aesop fable “The North Wind and the Sun”[26]), whilst the free-speech tasks required participants to speak at their usual volume and pace in response to two different questions (“What did you do last weekend?”; “What has your mood been like over the past couple of weeks?”), similar to the protocols used in[7, 27–29]. Self-report measures included three questionnaires covering demographics, current state (including questions relevant to voice quality at the time of recording, such as current physical and respiratory health, smoking/substance use, sleep, and device used), and health information (covering in depth questions relevant to chronic health conditions including diabetes, cardiovascular health, kidney disease, respiratory health, medication use, neurological/psychiatric health, and screening questions relevant to HbA1c testing).

#### 2.5.2 Stage 2: Remote HbA1c testing

HbA1c measures glycated hemoglobin, reflecting average blood glucose levels over the preceding three months, and is the gold-standard diagnostic test for T2D in the NHS. The test involved finger-prick blood collection (5ml) using a lancet, with results categorised as: *<*42 mmol/mol (6.0%): normal; 42-47 mmol/mol (6.0-6.4%): prediabetes; *≥*48 mmol/mol (6.5%): diabetes. Tests were sourced and processed via Medichecks Ltd., a United Kingdom Accreditation Service (UKAS)-accredited direct-to-consumer blood testing company.

To administer the HbA1c tests, participants were first divided into those with self-reported T2D and those without. Low-, medium- and high-risk groups among participants without self-reported T2D were then defined based on their predicted speech model scores, with individuals from across these groups recruited via Prolific to complete at-home HbA1c blood tests within three months of providing their voice data. High-risk and low-risk groups comprised those in the highest and lowest 10th percentile, respectively, while medium-risk participants were randomly selected from the intermediate score range. This sampling strategy resulted in a skewed distribution for HbA1c testing, with extreme risk scores overrepresented compared to the broader cohort that completed the self-report questionnaire. This stratified sampling strategy both ensured adequate representation across the predicted risk spectrum to assess discrimination between risk groups, and enriched for potential diabetes cases relative to random sampling, improving statistical power to evaluate model performance given the limited HbA1c testing resources.

HbA1c results provided objective, current diabetes status independent of self-reported diagnoses, accounting for individuals in remission or with undiagnosed diabetes. The continuous HbA1c values also enabled analysis across the spectrum of abnormal glycemic control, from prediabetes to diabetes, with thresholds set for analysis at HbA1c *≥* 42 mmol/mol or 6.0% and HbA1c *≥* 48 mmol/mol or 6.5%, respectively. For model validation, self-reported diabetes diagnoses from the health questionnaire were disregarded in favour of HbA1c-confirmed status, enabling assessment of the model’s ability to detect blood test-confirmed diabetes.

### 2.6 Statistical Analysis and Performance Evaluation

#### 2.6.1 Evaluation strategy

All performance evaluations were conducted using the reading out loud task only, with a single recording of 10 to 20s per participant. To obtain robust performance estimates with optimised decision thresholds, we employed a bootstrap cross-validation (CV) procedure. We created 1000 bootstrap resamples of the test set (N=7,319 participants), and for each bootstrap sample performed 3-fold cross-validation. Within each CV fold, the decision threshold was optimised on 2/3rd of the data (validation split) to maximize balanced accuracy, then performance metrics were calculated on the remaining 1/3rd (evaluation split). This yielded 3000 replicate performance measurements (1000 bootstrap iterations × 3 CV folds), allowing us to report mean ± standard deviation for all metrics. The primary outcomes of the speech-model included area under the curve (AUC), sensitivity (recall) and false positive rate (1 - specificity). We also evaluated Expected Calibration Error (ECE) using 10 equal-width bins to assess the alignment between predicted probabilities and observed accuracy across confidence levels.

#### 2.6.2 Comparison with QDiabetes

Model performance was compared against QDiabetes-2018, the NICE-recommended demographic risk assessment tool for diabetes screening in the UK[21]. QDiabetes calculates 10-year diabetes development risk based on age, BMI, ethnicity, family history, and clinical factors. While QDiabetes and our voice-based model address different clinical questions (future risk vs. current detection), QDiabetes functions as a case-finding tool in practice, and in the absence of any systematic non-invasive screening alternative, represents the closest available clinical benchmark. This comparison provides context for performance in population screening scenarios.

When comparing model performance, we use bootstrap significance testing[30, 31]. For each bootstrap sample, we compute the difference in the metric of interest (e.g., AUC) between models (speech-model minus QDiabetes), such that positive values indicate higher speech-model performance. This yields a distribution of differences; if the 95% confidence interval excludes 0, the difference is statistically significant. We also report a two-tailed p-value, calculated as twice the smaller proportion of bootstrap differences falling on either side of zero.

#### 2.6.3 Subgroup Analyses

To assess model generalisability and identify potential confounds, we evaluated performance across self-reported demographic subgroups: age (*≥* 40 or *<* 40), sex (male or female) and ethnicity (aggregated categories as recommended by the UK government[32]: White, Black, Asian, Mixed or Other, with Asian category further divided into South Asian and Other Asian to reflect the higher T2D risk in individuals of South Asian descent[33]); as well as clinical characteristics: BMI (*≥* 30 indicating obesity or *<* 30), comorbid conditions (CVD, hypertension, CKD and long COVID) and diabetes medication use (no medication or any medication). Unlike the full evaluation strategy (Section 2.6.1), subgroup analyses used AUC as the sole metric. Since AUC is threshold-independent, no threshold optimisation or cross-validation was required; bootstrap resampling (1000 iterations) was retained to estimate confidence intervals.

#### 2.6.4 Sensitivity to training label noise

To assess the impact of mixed-type diabetes training labels on model performance (i.e. type 1, type 2, gestational), we compared two speech models using 5-fold stratified cross-validation within the Stage 1 cohort (n = 7,319). The first model was trained on verified T2D labels collected in the present study. The second model replicated the model’s training conditions, using mixed-type diabetes labels and speech embeddings from an earlier data collection wave involving the same participants. Both models were evaluated against verified T2D status using the Stage 1 embeddings. Performance was compared using bootstrap resampling (1,000 iterations per fold, 5,000 total), with a two-tailed p-value computed as twice the minimum of the proportions of bootstrap samples where the AUC difference was less than or equal to zero or greater than or equal to zero.

## 3 Results

### 3.1 Participant Characteristics

Demographics and participant characteristics across the different stages of the study are outlined in Table 1.

**Table 1.** Participant Demographics and Characteristics Across Study Datasets. Demographic and clinical characteristics of participants in the training dataset (used for model development), self-report health dataset (Stage 1), and HbA1c dataset (Stage 2). To maximise data availability for model training, we utilised an existing dataset that employed broader, less granular labels for diabetes status (combining Type 1, Type 2, gestational, and other forms without distinction). Consequently, detailed demographic and clinical information collected in validation stages (such as specific ethnic groups, BMI and comorbidities) are not available for the training cohort.

| Datasets | Training data | Self-report health data (Phase 1) | HbA1c data (Phase 2) |
| --- | --- | --- | --- |
| **N** | 21,129 | 7,319 | 801 |
| **Diabetes prevalence** |  |  |  |
| Any | 1,318 (6.3%) | 467 (6.4%) | 35 (4.4%) |
| T2D | - | 217 (3.0%) | 35 (4.4%) |
| Confirmed | - | - | 29 (3.6%) |
| Undiagnosed | - | - | 6 (0.7%) |
| T1D | - | 58 (0.8%) | - |
| Gestational | - | 160 (2.2%) | - |
| Other (induced) | - | 32 (0.4%) | - |
| **# speech samples** | 63,283 | 7,319 | 801 |
| **Birth Sex** |  |  |  |
| Female | 13,341 (63.1%) | 4,917 (67.2%) | 496 (61.0%) |
| Male | 7,715 (36.5%) | 2,402 (32.8%) | 305 (38.1%) |
| Other | 73 (0.3%) | - | - |
| **Ethnicity** |  |  |  |
| White | 15,368 (72.7%) | 6,003 (82.0%) | 685 (85.5%) |
| Black | 3,079 (14.6%) | 547 (7.5%) | 45 (5.6%) |
| Asian | 1,109 (5.2%) | 476 (6.5%) | 46 (5.7%) |
| South Asian | - | 318 (4.3%) | 29 (3.6%) |
| Other Asian | - | 158 (2.2%) | 17 (2.1%) |
| Mixed | 1,107 (5.2%) | 244 (3.3%) | 20 (2.5%) |
| Other | 462 (2.2%) | 49 (0.7%) | 5 (0.6%) |
| **BMI** |  |  |  |
| <30 | - | 5,377 (35.4%) | 509 (31.1%) |
| ≥ 30.0 | - | 1,942 (26.5%) | 292 (36.5%) |
| **Age (years)** |  |  |  |
| < 40 | 12,162 (57.6%) | 3,970 (54.2%) | 312 (39.0%) |
| ≥ 40 | 8,967 (42.4%) | 3,349 (45.8%) | 489 (61.0%) |
| **Comorbidities** |  |  |  |
| CVD | - | 185 (2.5%) | 33 (4.1%) |
| Hypertension | - | 1,089 (14.9%) | 202 (25.1%) |
| CKD | - | 190 (2.6%) | 8 (1.0%) |
| **Medications** |  |  |  |
| No medication | - | 7,090 (96.9%) | 745 (93%) |
| Any diabetes medication | - | 229 (3.1%) | 56 (7.0%) |

Abbreviations: T2D = Type 2 diabetes; T1D = Type 1 diabetes; CVD = cardiovascular disease; CKD = chronic kidney disease; BMI = body mass index; Confirmed = self-reported cases of T2D; Undiagnosed = newly diagnosed cases of T2D in individuals who did not self-report as having T2D during Stage 1 data collection.

### 3.2 Model Performance

#### 3.2.1 Predicting T2D from self-reported health information

T2D detection performance based on self-reported data is described in Table 2. Assuming prediction models with AUC values above 0.80 can be considered clinically-useful[34, 35], we observed the speech-model was effective at distinguishing between people who self-reported a diagnosis of T2D (mean self-reported HbA1c level of 48.9*±*14.1 mmol/mol) and those who did not. The speech model also achieved an ECE of 0.019, usually considered to represent good calibration.

**Table 2.** Discrimination of T2D detection based on self report across speech and QDiabetes models. Values are shown as mean ± standard deviation across 3000 iterations.

| Metric | Speech model | QD |
| --- | --- | --- |
| AUC | $0.80 \pm 0.03$ | $0.86 \pm 0.03$ |
| Recall (Sensitivity) | $0.76 \pm 0.10$ | $0.82 \pm 0.07$ |
| False Positive Rate (1 - Specificity) | $0.31 \pm 0.06$ | $0.24 \pm 0.03$ |
| ECE | $0.019 \pm 0.004$ | $0.017 \pm 0.004$ |

Abbreviations: AUC = Area Under Receiver Operating Characteristic Curve; ECE = Expected Calibration Error; QD = QDiabetes.

#### 3.2.2 Speech model vs QDiabetes for predicting self-reported diabetes

For overall prediction of T2D, the speech model showed modestly lower overall discrimination than QDiabetes (ΔAUC = *−*0.06, 95% CI [*−*0.08, *−*0.03], *p <* 0.001; Table 2, Figure 2). Differences in sensitivity (*p* = 0.29) and specificity (*p* = 0.11) were not statistically significant. Calibration was good for both models (ECE of 0.019 and 0.017), with no statistical difference (p=0.15). Similarly, both models discriminated well between self-reported T2D and type 1 diabetes, with no statistically significant difference in AUC (ΔAUC = *−*0.02, 95% CI [*−*0.09, 0.05], *p* = 0.54; Table 3). However, when extending to all past diabetes diagnoses (including gestational and other induced forms), the speech model significantly outperformed QDiabetes (ΔAUC = 0.17, 95% CI [0.12, 0.22], *p <* 0.001), with QDiabetes discrimination dropping substantially (AUC = 0.62) while the speech model remained stable (AUC = 0.79).

**Fig. 2.**
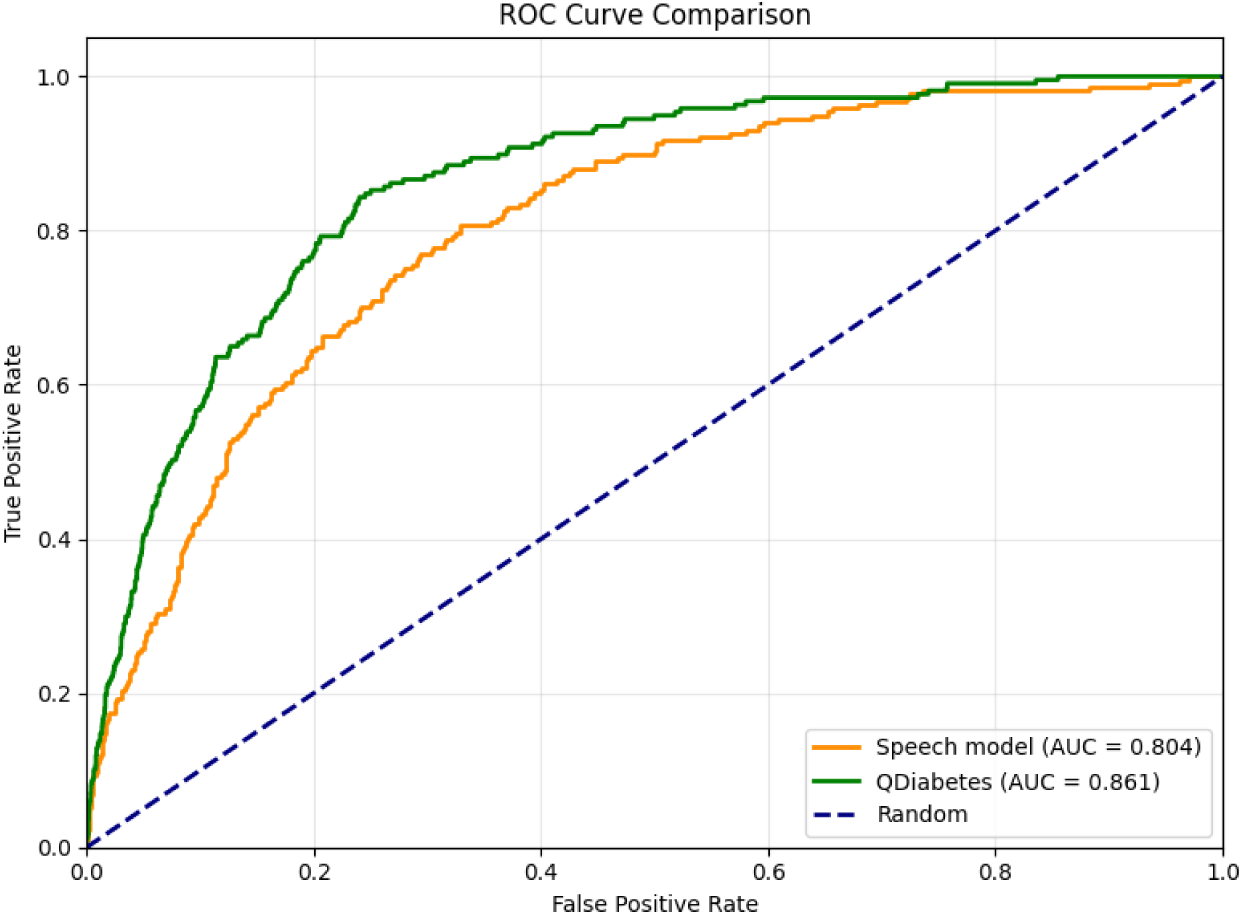
ROC curves comparing the speech model and QDiabetes for T2D prediction on self-reported health data (Stage 1).

**Table 3.** Performance of the speech model and QDiabetes when discriminating T2D from self-reported type 1 diabetes and any present or past diabetes diagnosis (including type 1, gestational and other induced forms of diabetes).

| Subgroup/Condition | Sample size (T2D/sub-group) | Prevalence | Speech model AUC | QD AUC |
| --- | --- | --- | --- | --- |
| Overall | 217/7319 | 3.0% | 0.80 | 0.86 |
| On population with: |  |  |  |  |
| Type 1 + Type 2 diabetes only | 217/275 | 79.0% | 0.81 | 0.83 |
| Any diabetes diagnosis | 217/467 | 46.5% | 0.79 | 0.62 |

Abbreviations: AUC = Area Under Receiver Operating Characteristic Curve; T2D = Type 2 Diabetes; QD = QDiabetes.

#### 3.2.3 Speech model’s performance across demographic subgroups

T2D prevalence varies substantially across demographic groups and frequently co-occurs with conditions that may themselves affect voice characteristics. To evaluate model’s robustness and identify potential sources of bias or confounding, we assessed its T2D prediction performance across demographic subgroups (birth sex, age and ethnicity), chronic comorbid conditions and medication status (Table 4).

**Table 4.** Overview of type 2 diabetes (T2D) in overall sample and common comorbid health condition and medication subgroups, speech-model and QDiabetes predictions also listed. ΔAUC = speech-model AUC *−* QDiabetes AUC. Positive values indicate higher speech model performance.

| | Sample size (T2D /sub-group size) | T2D prevalence (%) | Speech model AUC | QD AUC | $\Delta\text{AUC}$(95% CI) | $p$ |
| --- | --- | --- | --- | --- | --- | --- |
| **Overall** | 217/7319 | 3.0% | 0.80 | 0.86 | −0.06 (−0.09, −0.03) | <0.001 |
| **With comorbidities:** |  |  |  |  |  |  |
| CVD | 21/185 | 11.4% | 0.69 | 0.77 | −0.08 (−0.22, 0.05) | 0.204 |
| Hypertension | 114/1090 | 10.4% | 0.65 | 0.72 | −0.07 (−0.13, −0.01) | 0.018 |
| Obesity (BMI $\geq 30$) | 125/1910 | 6.6% | 0.73 | 0.80 | −0.07 (−0.12, −0.03) | <0.001 |
| CKD | 18/190 | 9.4% | 0.82 | 0.83 | −0.01 (−0.08, 0.07) | 0.908 |
| Long COVID | 10/121 | 8.3% | 0.74 | 0.59 | 0.15 (−0.01, 0.31) | 0.054 |
| **Without comorbidities:** |  |  |  |  |  |  |
| No CVD | 196/7134 | 2.8% | 0.80 | 0.86 | −0.06 (−0.09, −0.03) | <0.001 |
| No hypertension | 103/6230 | 1.6% | 0.83 | 0.86 | −0.03 (−0.06, 0.01) | 0.12 |
| No obesity (BMI < 30) | 91/5333 | 1.7% | 0.81 | 0.86 | −0.05 (−0.09, −0.003) | 0.04 |
| No CKD | 199/7129 | 2.8% | 0.80 | 0.86 | −0.06 (−0.09, −0.03) | <0.001 |
| No long COVID | 207/7199 | 2.9% | 0.81 | 0.87 | −0.06 (−0.09, −0.04) | <0.001 |
| **By medication use:** |  |  |  |  |  |  |
| No medication | 64/7090 | 0.9% | 0.74 | 0.80 | −0.06 (−0.12, 0.003) | 0.034 |
| Any medication | 153/229 | 66.8% | 0.81 | 0.81 | 0.00 (−0.06, 0.07) | 0.866 |

Abbreviations: AUC = Area Under Receiver Operating Characteristic Curve, CKD = Chronic Kidney Disease; QD = QDiabetes, BMI = Body Mass Index.

The model demonstrated robust performance across most demographic subgroups (see Figure 3). Sex-specific performance was comparable (male AUC = 0.79; female AUC = 0.81), as was performance across age categories (AUC *≥* 0.75). Ethnicity-specific performance remained strong for most subgroups, including participants who reported belonging to White, South Asian, Mixed and Other ethnic groups (AUC *≥* 0.8), with lower performance observed in Black (AUC = 0.69) and Other Asian (AUC = 0.65) participants. However, the latter two subgroups had limited representation of T2D cases (*n* = 18 and *n* = 5, respectively), despite reasonable overall sample sizes (*n* = 547 and *n* = 159), which limits the reliability of these findings.

**Fig. 3.**
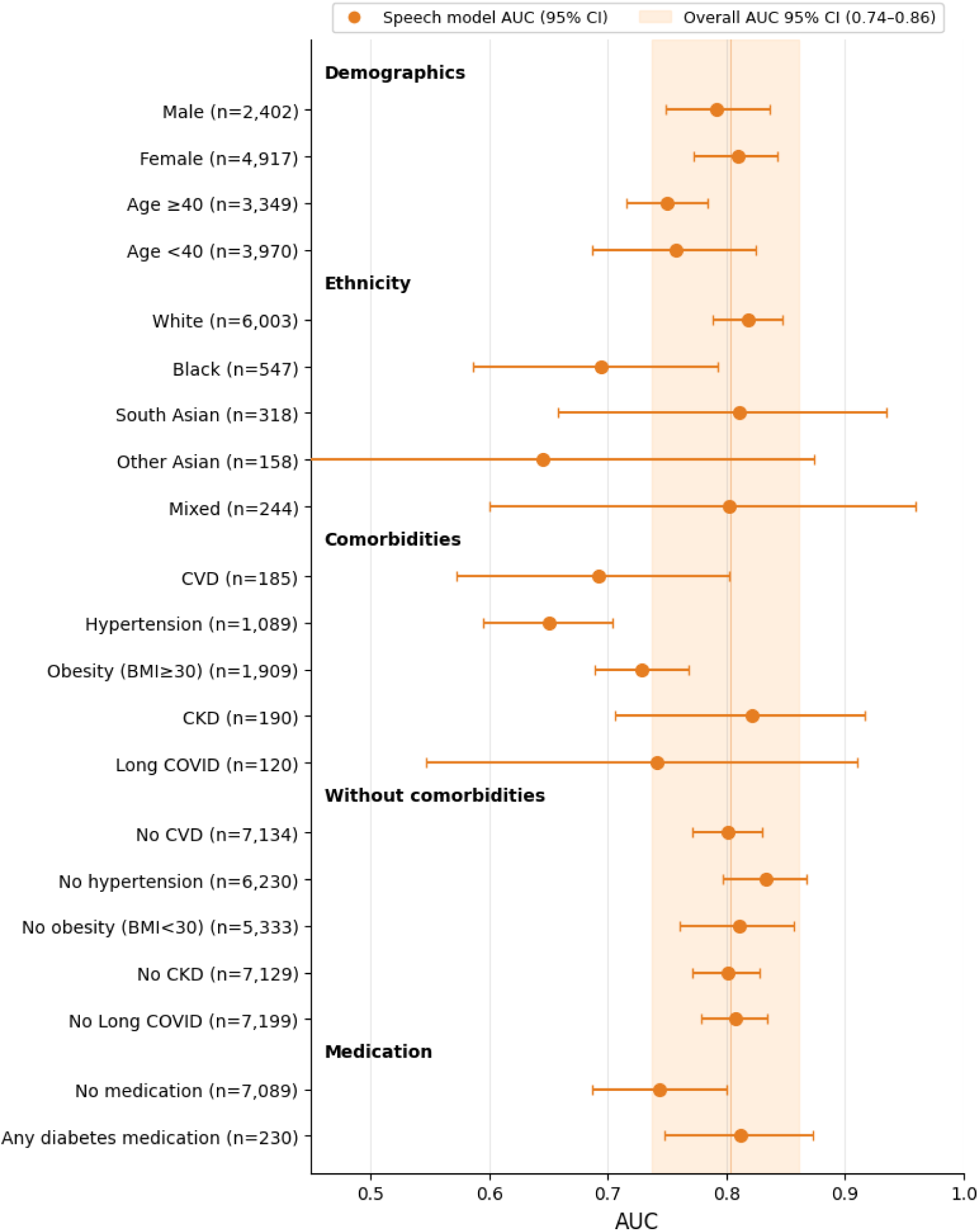
Area Under the Curve (AUC) and 95% Confidence Interval (CI) when predicting Type 2 Diabetes (T2D) in different subgroups. CVD = cardiovascular disease, CKD = chronic kidney disease, BMI = body mass index.

#### 3.2.4 Speech model’s performance across comorbidities and medication status

T2D is often co-morbid with multiple other chronic health conditions, including CVD, hypertension, BMI *≥* 30 indicating obesity, and CKD. We observed lower performance predicting T2D in groups with most of the conditions that commonly co-occur with T2D (CVD AUC = 0.69; hypertension AUC = 0.65; BMI *≥* 30 AUC = 0.73) but comparable performance for CKD (AUC = 0.82) and other groups without these respective conditions (AUC *≥* 0.8; Figure 3). A similar pattern was observed for QDiabetes, which also showed reduced performance in comorbid groups while maintaining strong discrimination in non-comorbid populations (Table 4).

We also examined performance in individuals with long COVID, a condition increasingly linked to T2D[36]. Here, the speech model retained reasonable discrimination (AUC = 0.74) whereas QDiabetes performance dropped substantially (AUC = 0.59), though the difference did not reach statistical significance (ΔAUC = 0.15, 95% CI [*−*0.005, 0.31], *p* = 0.054; Table 4).

Finally, we examined how diabetes medication use affects model performance. We found that both models maintain an AUC of 0.81 within people taking at least one type of diabetes medication (*N* = 229 of which 66.8% have T2D), and drops slightly for people taking no diabetes medication (*N* = 7, 090 of which 0.9% have T2D) for the speech model (AUC = 0.74) but stayed constant for QDiabetes (AUC = 0.80, ΔAUC =*−*0.06, 95% CI [*−*0.119, *−*0.003], *p* = 0.034).

#### 3.2.5 Speech model validation using HbA1c across diabetes severity thresholds

At the diabetes threshold (HbA1c *≥* 48 mmol/mol or 6.5%, with mean HbA1c level of 60.7*±*13.3 mmol/mol), the speech model achieved an AUC of 0.75 (*±*0.07), correctly identifying 82% (*±*23%) of diabetes cases, though 47% (*±*11%) of non-diabetic individuals were classified as high-risk. By comparison, QDiabetes demonstrated comparable performance in the same cohort (AUC 0.77*±*0.08) with a 72% (*±*22%) sensitivity and 32% (*±*12%) false positives. No differences reached statistical significance for any metric (ΔAUC = *−*0.02, 95% CI [*−*0.11, 0.05], *p* = 0.60; sensitivity *p* = 0.51; specificity *p* = 0.15).

To check if the lower AUCs as compared to self-reported diabetes were due to the skewed population distribution in the reduced HbA1c evaluation set (801 participants vs 7,319 for self-reported data), which was selected based on predicted risk groups, as described in 2.5.2., we also computed AUC for the speech model and QDiabetes for this reduced population using the self-reported label. We found AUCs of 0.72*±*0.06 and 0.76*±*0.06, respectively, comparable to the results obtained above, and with no significant differences for any metric (ΔAUC = *−*0.04, 95% CI [*−*0.09, 0.02], *p* = 0.20). This confirms that the lower AUCs observed for HbA1c-based diagnosis are attributable to the skewed population distribution in the reduced HbA1c evaluation set, rather than to a degradation in model discriminative performance.

At the prediabetes threshold (HbA1c *≥* 42 mmol/mol or 6%, with mean HbA1c level of 53.9*±*12.9 mmol/mol), the model achieved AUC 0.73 (*±*0.06), detecting 79% (*±*16%) of cases with 43% (*±*9%) false positives, while QDiabetes demonstrated stronger discrimination at this threshold (AUC 0.80 *±* 0.06, 79 *±* 13% sensitivity, 30 *±* 7% false positives). Again, the difference in AUC was not statistically significant (ΔAUC = *−*0.06, 95% CI [*−*0.13, 0.00], *p* = 0.054), and the recall and specificity differences were also non-significant (*p* = 0.97 and *p* = 0.09, respectively).

#### 3.2.6 HbA1c discrimination between predicted risk groups in undiagnosed population

The model effectively distinguished between individuals at different levels of diabetes risk, even within the undiagnosed (no self-reported T2D) sample (Figure 4). Participants predicted to be at high risk of undiagnosed diabetes (top 10th percentile; *n* = 175) had significantly higher mean HbA1c levels than both the medium-risk group (intermediate score range; *n* = 478; Δmean = +3.3 mmol/mol, *p <* 0.001; Mann-Whitney U tests with Bonferroni correction for three pairwise comparisons) and the low-risk group (bottom 10th percentile; *n* = 83; Δmean = +5.3 mmol/mol, *p <* 0.001). Conversely, low-risk participants had significantly lower HbA1c levels than the medium-risk group (Δmean = *−*2.1 mmol/mol, *p <* 0.001). Importantly, none of the participants in the low-risk group had HbA1c levels in the prediabetic (*≥* 42 mmol/mol) or diabetic (*≥* 48 mmol/mol) range, confirming the model’s ability to accurately identify individuals unlikely to have elevated blood glucose.

**Fig. 4.**
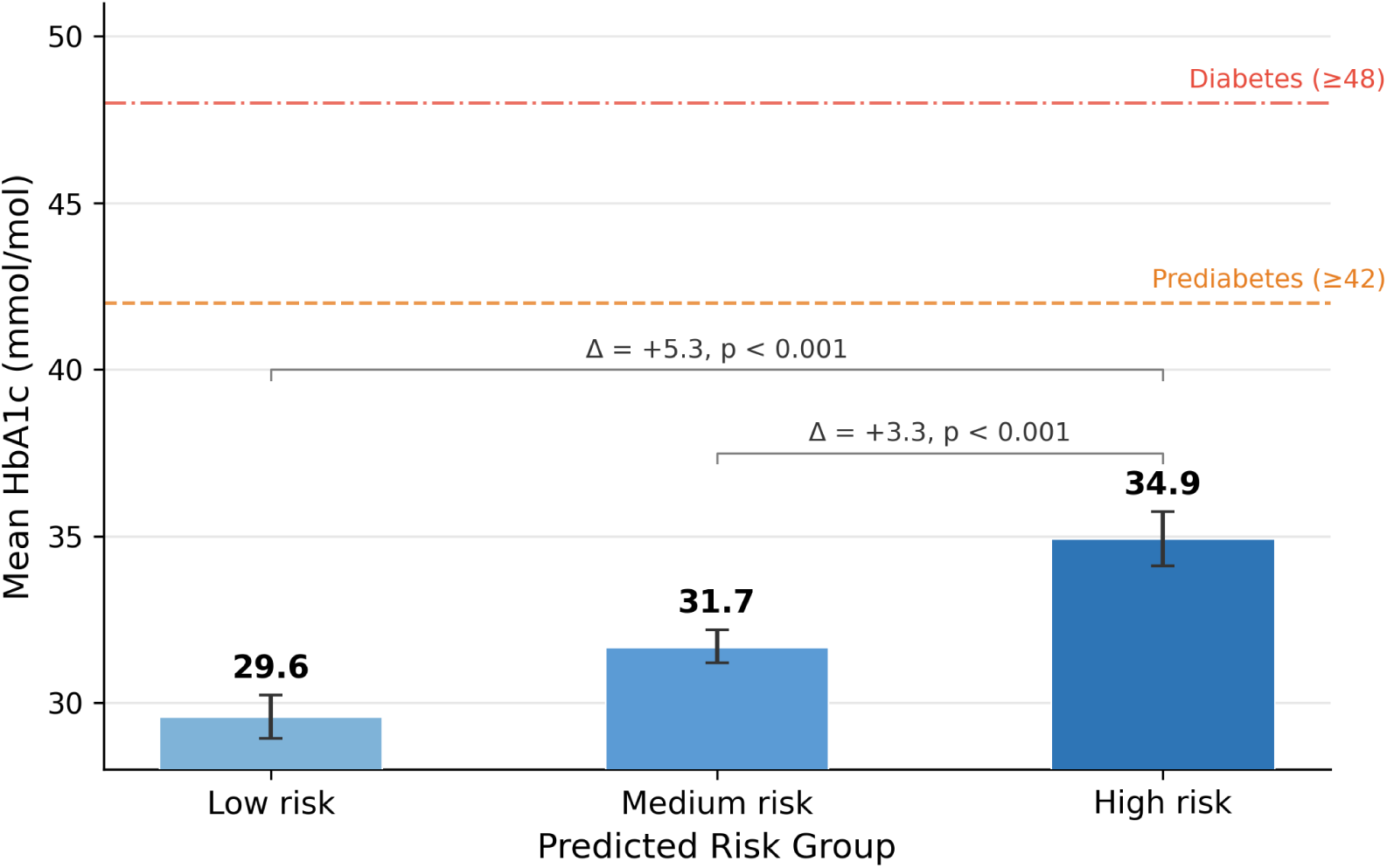
Mean HbA1c levels for different model-predicted risk groups (low, medium and high). Undiagnosed population only (no self-reported T2D, n=736)

#### 3.2.7 Sensitivity to training label noise

To assess sensitivity to the mixed-type diabetes labels used for model training, we retrained the model using 5-fold cross-validation within the Stage 1 cohort with clean T2D labels (*n* = 7, 319; 217 T2D cases), as well as again with the mixed-type label used in training, as described in Section 2.6.4. The clean-label model achieved AUC 0.775, compared to 0.740 for the mixed-label model (ΔAUC = 0.035, 95% CI [*−*0.038, 0.113], *p* = 0.34). The difference was not statistically significant, suggesting the mixed-type training labels did not substantially compromise T2D discrimination. Nevertheless, the trend toward higher AUC with clean T2D labels suggests that training on a larger dataset with verified T2D diagnoses could yield meaningful improvements, as the models are trained here with only *∼*5,800 speakers per fold.

## 4 Discussion

This study addressed three critical issues in speech-based diabetes detection: the need for large-scale validation, clinical biomarker validation and comparison to current recommended screening methods. By conducting a two-stage validation study in 7,319 UK adults, a much larger and more diverse sample than prior validation studies[13–15], we demonstrated that machine learning analysis of 20-second speech recordings can reliably distinguish individuals with T2D from those without.

The speech model, trained on the largest real-world sample to date, achieved an AUC of 0.80 for discriminating between individuals with and without T2D. This is on par with the 0.80 threshold considered clinically useful[37], though we acknowledge such thresholds lack standardised justification[34]. The model was well calibrated (ECE = 0.019), ensuring that predicted probabilities closely correspond to observed rates of T2D in the validation cohort. Performance approached that of QDiabetes (AUC = 0.86), the risk assessment tool recommended by NICE for the NHS T2D risk identification programme[21]. In the absence of a system-wide non-invasive screening pathway, T2D detection in the UK being largely opportunistic and reliant on blood testing, QDiabetes represents the most relevant available benchmark. Risk prediction algorithms like QDiabetes, while nominally predicting future T2D incidence (in this case, 10-year risk), often function as case-finding tools in practice. We therefore evaluated both tools on the same task, identifying current T2D status, providing a direct performance comparison[20]. At optimised decision thresholds, our model correctly identified three-quarters of T2D cases (sensitivity 0.76), with a false positive rate of approximately 30%. No significant differences were found for sensitivity (*p* = 0.29) or specificity (*p* = 0.11) between the speech model and QDiabetes, indicating that despite lower overall discrimination, the two methods performed comparably at their respective operating points. This suggests voice-based screening could serve a role in diabetes detection pathways, offering a non-invasive, fast and scalable first-line option for risk prediction that can be followed up with more time- and resource-intensive definitive blood tests if risk is identified.

An unexpected yet valuable finding emerged when evaluating broader diabetes categories: We examined a subset of participants who reported any diabetes diagnosis (past or present, including type 1, gestational, or drug-induced forms) and evaluated how well the model distinguished T2D from these other diabetes types. Voice-based predictions remained consistent, while QDiabetes performance dropped substantially. This may be because other forms of diabetes (often temporary, e.g., gestational diabetes), share risk factor profiles with T2D, for example, the demographics and clinical history of affected individuals[38], and QDiabetes cannot discriminate well between these overlapping characteristics. In contrast, the speech model captures paralinguistic features that may reflect both demographic characteristics embedded in voice and pathophysiological changes associated with poor glycemic control, potentially enabling more precise discrimination between current and past cases.

Another interesting exploratory finding was that, in contrast to QDiabetes (AUC = 0.59), the speech model retained reasonable performance in individuals with long COVID (AUC = 0.74). COVID-19 has been associated with a disproportionate increase in T2D cases[39]. In turn, T2D is itself a risk factor for developing long COVID, with the two conditions also sharing several important comorbidities, including cardiovascular and kidney disease[36]. QDiabetes’ reliance on predefined risk factors, established through historical population statistics and updated infrequently, may limit its adaptability to emerging conditions like long COVID that alter diabetes risk profiles. In contrast, data-driven models, such as the speech model used in this study, can be readily implemented to discover patterns directly from data without requiring pre-specification of risk factors. This creates potential advantages when new health conditions emerge that affect diabetes risk, or population risk profiles shift over time. Nevertheless, given the modest sample size of long COVID cases (n = 121), these findings remain preliminary and warrant validation in larger, dedicated studies.

Stratified analyses across demographic and clinical subgroups revealed generally consistent performance. Sex-specific discrimination was comparable (male AUC 0.79; female AUC 0.81) and also exceeded the gender-specific results reported by an earlier voice-biomarker diabetes screening model who achieved AUC values of 0.75 and 0.71, respectively[14]. Performance also remained robust across most age categories (AUC *≥* 0.75) and ethnic groups (AUC *>* 0.80). However, discrimination declined among Black and Other Asian participants (AUC *≤* 0.70). This warrants further investigation using samples with higher representation of T2D cases (*n* = 18 and *n* = 5 T2D cases in the current samples of *n* = 547 Black and *n* = 159 Other Asian individuals, respectively), both because of the higher diabetes burden in these ethnic groups[1] and the purported differences in T2D-related voice characteristics across ethnicities[18] (although see[40]).

The speech model performed worse among samples with CVD (AUC = 0.69), hypertension (AUC = 0.65) and obesity (BMI *≥* 30, AUC = 0.73). The reduced discrimination in individuals with these conditions likely reflects substantial physiological overlap. For example, both obesity and T2D share common pathophysiological mechanisms—insulin resistance, chronic inflammation, and metabolic dysfunction[41]—that may produce similar vocal changes. Additionally, obesity directly affects voice, such as through increased adipose tissue in the neck and chest, altering respiratory dynamics and vocal tract resonance[42]. Similarly, vocal changes have been observed in individuals with hypertension, which, if untreated, may progress to cardiovascular disease, although the underlying mechanisms in this case are less clear[43, 44]. From a screening perspective, this limitation is not necessarily problematic. CVD, hypertension and obesity are strongly associated with undiagnosed T2D prevalence and represent priority populations for screening[45]. A tool that flags these individuals for confirmatory blood glucose testing, even if the vocal signatures partially reflect comorbid pathophysiology rather than T2D-specific vocal changes, still achieves the clinical objective of identifying individuals who warrant further evaluation for potential undiagnosed disease.

Surprisingly, we did not observe a decline in the speech model’s performance when predicting T2D in individuals with CKD (AUC 0.82), another chronic condition associated with T2D. CKD often develops over time in poorly controlled T2D and is exacerbated by hypertension. Given that the speech models ability to predict T2D in this group is preserved or even slightly better, it may be that vocal changes associated with CKD are distinct from those seen in T2D. Alternatively, CKD as a result of current T2D may result in vocal changes that are informative for the model.

The performance pattern observed in comorbid populations suggests the speech model is not simply exploiting comorbidities as predictive shortcuts. If that were the case, we would expect substantially worse performance in their absence. However, the model maintains strong discrimination ability (*AUC ≥* 0.80) in non-comorbid populations, indicating that comorbidity status is not the primary driver of predictive performance. Notably, AUC never falls below 0.65 in any subgroup, indicating the model retains meaningful above chance discriminative ability across the full range of clinical presentations.

It is not well understood whether diabetes medication affects voice, inadvertently helping the speech model predict individuals with T2D. We therefore evaluated how well the model performed in individuals on any diabetes medication as compared to unmedicated participants. We found that prediction performance in the medicated group (n = 229) was similar to that in the overall sample (AUC 0.81). In contrast, in the group taking no diabetes medication (n = 7090), performance dropped modestly to an AUC of 0.74. Given that the complexity of diabetes therapy correlates with disease severity[46], this difference likely reflects a harder discrimination problem rather than a medication-specific vocal effect: unmedicated individuals may disproportionately represent milder disease, where T2D-associated physiological changes in speech are less salient. Conversely, the medicated group likely comprises individuals with more active and severe disease, consistent with the model’s capacity to detect pathophysiological signals across the continuum of disease severity.

For Stage 2 HbA1c validation, we tested a subset of 801 participants drawn from the original sample of 7,319 using home blood test kits supplied by a UK-accredited direct-to-consumer blood test company. To our knowledge, this is the only large-scale speech-based diabetes study to incorporate concurrent remote blood sampling within three months of voice recording. The use of HbA1c tests has confirmed that the speech model can detect physiological changes associated with diabetes across clinically relevant biomarker thresholds, with consistent performance in both diabetes (HbA1c *≥* 48 mmol/mol or 6.5%: AUC 0.75) and prediabetes (HbA1c *≥* 42 mmol/mol or 6.0%: AUC 0.73). While performance was lower than on self-reported data, a comparable reduction was observed for QDiabetes (AUC 0.77 and 0.80, respectively), suggesting this reflects the distributional characteristics of the reduced HbA1c subsample rather than a fundamental limitation of either model.

The relatively similar performance of the speech model across both diabetes thresholds (diabetic and prediabetic) is notable given that vocal changes associated with metabolic dysregulation are expected to become more pronounced with disease progression. HbA1c reflects average blood glucose levels over the preceding three months and may not reflect levels at the time of the speech recording. However, it is possible that paralinguistic aspects of speech picked up by the model vary with the level of blood glucose, irrespective of the disease stage[47]. Several studies have also proposed that it is the variability in blood glucose levels, rather than elevated blood glucose itself, that causes detrimental effects in diabetes, including oxidative stress, neuropathy and retinopathy, more common in advanced disease stages[48–50]. Future studies that measure blood glucose variability continuously could help further characterize how the speech model’s paralinguistic features contribute to its predictions. Although confound analyses suggest the model is largely robust, residual demographic signal cannot be excluded. This would not diminish screening utility, as demographic characteristics are well-established predictors of diabetes risk. The ability to infer them passively from short segments of speech, without requiring explicit self-report, represents a practical advantage in populations where competing priorities and limited access to GP services contribute to low preventive screening uptake[2].

Finally, the model demonstrated good discriminative performance in individuals at different levels of diabetes risk within the sample with no self-reported diabetes diagnosis. Participants predicted to be at high risk of T2D had significantly higher mean HbA1c levels as compared to medium- or low-risk groups. Furthermore, none of the low-risk participants, as predicted by the model, had concerning levels of HbA1c. This finding underscores the potential of speech-based screening as a tool for population-wide diabetes risk stratification.

Overall, the findings of this study suggest that voice-based tools could be used for diabetes screening. We present a model that was trained on the largest real-world dataset to date and which achieved clinically useful discrimination on self-reported data (AUC = 0.80). Importantly, it maintained strong performance when validated against HbA1c biomarkers (AUC = 0.75, AUC = 0.73), where both diabetic and non-diabetic labels were objectively confirmed through blood testing. This demonstrates the model’s ability to detect diabetes beyond known, self-reported cases, addressing a critical gap given that approximately 1 million individuals in the UK remain undiagnosed.

The practical advantages of voice-based screening extend beyond predictive performance (see Figure 5). Current screening pathways in the UK rely either on opportunistic blood tests taken during regular GP visits, or time-intensive screening appointments, such as the NHS Health Check, which require 20-30 minutes and include blood draws for glucose and lipids, with approximately 40% of eligible individuals not attending[3]. In contrast, speech-based screening requires only a 20-second recording obtainable remotely without needles, clinical visits, questionnaire completion or recall of detailed medical history. It is not disease-specific and may enable multiplex screening by flagging the risk of several diseases at the same time (e.g., hypertension[44], respiratory conditions[51, 52] or depression and anxiety[7]), maximising clinical yield while maintaining low burden. Furthermore, speech-based approaches are inherently scalable and compatible with both digital and telephone platforms. This could reduce barriers to participation for individuals who face challenges accessing conventional healthcare, whether due to aversion to blood tests, time constraints, mobility limitations, or geographic distance from clinical facilities[2]. Only those individuals who are flagged to be at high risk could be asked to complete blood testing to confirm diagnosis and administer lifestyle or clinical interventions, optimising diabetes screening and saving patient and clinician time. The technology’s minimal friction may particularly appeal to younger, digitally-native populations who are often underrepresented in preventive screening but will age into higher-risk categories. In addition, if done correctly, for example ensuring representation in both model training and tool design[53, 54], the implementation of such technology has the potential to close the gap in health inequities[1]. Therefore, we suggest that future versions of our model may meaningfully complement existing diabetes detection strategies.

**Fig. 5.**
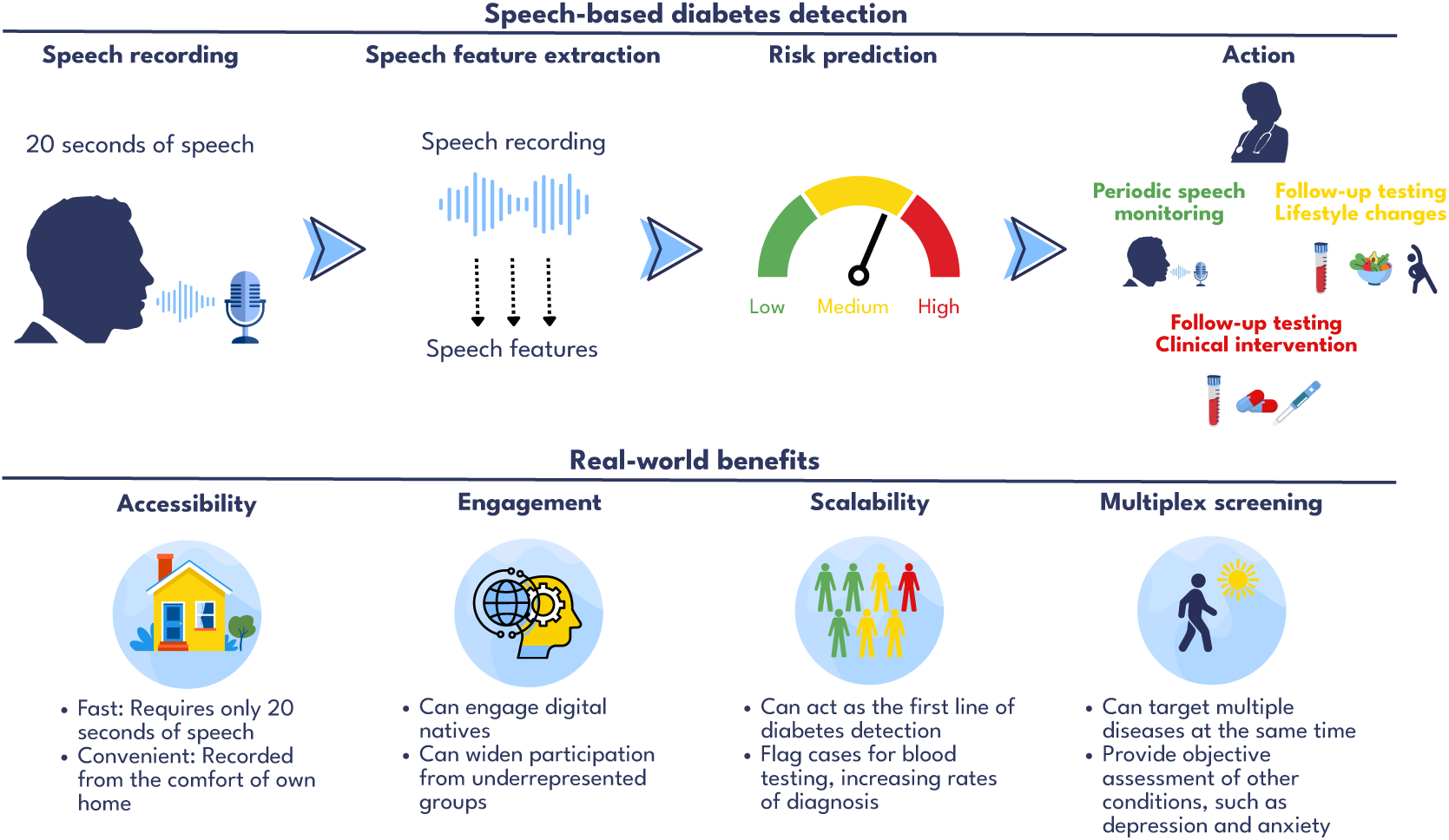
Summary of what speech-based diabetes detection could look like and real-world benefits of this approach.

In practice, voice-based tools may be implemented as the first line of T2D screening (see Figure 5). Speech recordings can be made before (e.g., as a stand-alone screening) or during primary care appointments, either on the telephone, digitally or in person. Features automatically extracted from these recordings can be passed on to the risk prediction model to generate a risk score. The latter can then be used by the primary care practitioners to triage patients. Low-risk patients could be asked to come back for periodic speech-based screening. Individuals with medium risk scores could be invited for confirmatory blood testing and be given access to a digital lifestyle intervention programme to develop healthier diet and exercise habits. Finally, everyone identified as high-risk, would also receive blood workup to confirm diagnosis, followed by a clinical intervention. This tiered approach would reduce reliance on time-consuming demographic questionnaires and invasive blood testing where they are not warranted, generating cost savings and releasing clinician time for direct patient care. In summary, this study presents a large-scale, biomarker-validated speech-based screening tool for T2D. Using a cohort twelve times larger than any previous comparable studies, we demonstrated that 20-second voice recordings can detect diabetes with discrimination approaching QDiabetes (AUC = 0.80 vs 0.86), the NICE-recommended but heavily underutilised screening tool for UK primary care[21], and the most relevant available benchmark in the absence of any systematic non-invasive screening alternative. Our speech model was robust to most demographic confounds. The HbA1c validation provided preliminary evidence that this model can identify T2D cases beyond self-reported diagnoses, including in individuals with undiagnosed disease. Its minimal burden, requiring no clinical appointments, specialised personnel, or invasive procedures, positions speech-based screening as a scalable and practical complement to existing detection pathways. Combined with its potential for multiplex screening from a single speech sample, and future work on improving performance in ethnic minorities and in the presence of comorbidities, this approach could meaningfully expand access to preventive healthcare, particularly for populations facing barriers to conventional screening.

## Acknowledgements

We thank all participants who contributed to this study. We are also grateful to the Medichecks team for their support. We acknowledge the contributions of consultant physicians Dr Joanna Bilak (MBBS MRCP) and Dr Andrew Solomon (BM BCh MA DM FRCP), and Paula Mason (RD, CDCES; registered dietitian and diabetes educator) for their consultation on the health questionnaire and experimental design.

## Author contributions

S.G., E.M. and E.B conceived the study, E.B. and A.L.G. conducted the study, R.P., E.B. and O.P. analysed the results. R.P., E.B., O.P., A.L.G, G.Č., B.d.C., S.G. and E.M. contributed to the writing and editing of the manuscript.

## Data availability

The datasets generated and analysed during the current study are not publicly available due to lack of consent for public sharing of raw data, which due to the nature of speech data would compromise participant privacy. They may be made available on reasonable request to the corresponding author for non-commercial research purposes related to detecting or monitoring diabetes (the purposes for which consent for research data sharing was obtained), subject to completion of a Data Sharing Agreement with thymia Ltd.

